# Decreased within-default mode network connectivity with accelerated intermittent theta burst stimulation across unipolar and bipolar depression: Candidate transdiagnostic circuit marker for treatment response

**DOI:** 10.64898/2025.12.24.25342970

**Authors:** Yvette I. Sheline, Alexandra S. Batzdorf, Walid Makhoul, Nicholas L. Balderston, Kevin G. Lynch, Robin F. H. Cash, Conor M. Liston

## Abstract

**Background:** An accelerated schedule of intermittent theta burst stimulation (aiTBS) has been shown to improve clinical efficacy of transcranial magnetic stimulation for bipolar disorder (BD) and major depressive disorder (MDD). We investigated the functional connectivity changes underpinning clinical effects of aiTBS on transdiagnostic treatment-resistant depression (TRD).

**Methods:** Data were collected from two studies: an open-label active aiTBS study for MDD and a double-blind, randomized control aiTBS study for BD. Thirty-four participants (18 female/16 male) with transdiagnostic TRD currently experiencing moderate to severe depressive episodes (Montgomery-Åsberg Rating Scale [MADRS] score ≥ 20). Participants received active (22 total; 12 BD/10MDD) or sham (12 BD) aiTBS delivered to personalized left dorsolateral prefrontal cortical targets. Participants completed MRI scans before and after ten sessions of active or sham aiTBS per day for five days. Within each aiTBS group (active/sham), connectivity changes within the default mode network (DMN) following aiTBS and their correlations with MADRS scores were evaluated.

**Results:** Immediately following aiTBS, active group members had lower within-DMN connectivity (*p* = .01, *d* = –0.62) and lower MADRS scores (*p* < .0001) than at baseline; these changes were correlated (*r* = .47, *p* = .03). They also reported decreased insomnia (*p* < .0001) and suicidality (*p* < .0001). These changes were not present in the sham BD group (*p*s = .29– .92).

**Conclusions:** aiTBS was associated with decreased within-DMN connectivity among people with transdiagnostic TRD, which could underlie depression improvement.

The study is registered on ClinicalTrials.gov (study identifier NCT05228457). https://clinicaltrials.gov/study/NCT05228457

## INTRODUCTION

Treatment-resistant depression (TRD) afflicts approximately 30% of individuals with major depressive disorder (MDD) and bipolar disorder (BD), contributing to elevated suffering, functional impairment, and costs up to 19 times higher than for treatment responders (Crown et al., 2002; Fava & Davidson, 1996). Depression-related dysfunction is increasingly conceptualized as arising from disruptions in distributed neural networks rather than specific regional locations (Fornito et al., 2015). Meta-analyses have shown inconsistent region of interest (ROI) findings across transcranial magnetic stimulation (TMS) depression studies. For example, a meta-analysis examining activation abnormalities in ROI data derived from > 1000 patients across 57 studies and 99 individual neuroimaging experiments demonstrated a lack of significant anatomical convergence, highlighting an absence of consensus (Muller et al., 2017). In contrast, a network based approach (Cash et al., 2023) identified brain circuits—rather than isolated regions—underlying emotional and cognitive dysfunction in MDD. These circuits align with lesion-based depression networks and effective TMS/deep brain stimulation targets. This recent meta-analysis across 1058 individual neuroanatomical foci from emotional and cognitive task-based experiments in MDD (Cash et al., 2023) identified consistent, robust alterations in large-scale networks, including the dorsomedial prefrontal region of the default mode network (DMN), despite neuroanatomical heterogeneity. Accordingly, localization of psychiatric symptoms, including in the context of depression, has increasingly shifted focus from individual brain regions to distributed brain networks (Downar & Daskalakis, 2013).

A challenge in treating depression is the relatively slow onset of symptom improvement. Transcranial magnetic stimulation (TMS) can be an effective therapy for TRD, but conventional TMS therapy involves four to six weeks of daily treatment. In contrast, accelerated intermittent theta-burst stimulation (aiTBS) condenses the TMS treatment course into multiple treatments per day to achieve a more rapid, durable clinical response (Cole et al., 2020; Holtzheimer et al., 2010). Some forms of aiTBS have resulted in rapid remission of depressive symptoms in MDD (Li et al., 2014; Rachid, 2019), leading to FDA approval. More recently, we demonstrated rapid remission of depressive symptoms in BD with aiTBS (Sheline et al., 2024). While effects of antidepressant treatment on clinical outcome have been described, relatively little is known about brain functional correlates. In the current study, we used a circuit-based approach to investigate change in within-DMN connectivity following aiTBS in a transdiagnostic cohort of treatment-resistant individuals with MDD and BD using the same stimulation procedures and clinical assessment instruments across diagnostic samples. Our aiTBS protocol utilized a high pulse number and stimulation targets were optimized to align with individual-specific network architecture. Our goal was to determine whether within-DMN connectivity would be affected by aiTBS treatment similarly across the transdiagnostic cohort and whether connectivity changes would coincide with depressive symptom improvement. This focus was motivated by our prior work, and that of others, highlighting the importance of the DMN in depression pathophysiology (Hamilton et al., 2015; Sheline et al., 2009; Sheline et al., 2010) and its treatment using TMS (Liston et al., 2014). This approach differs from prior studies by examining within-DMN connectivity across both MDD and BD to determine whether these transdiagnostic changes were associated with decreases in depression symptomatology.

## METHODS AND MATERIALS

### Study Design

The BD sample was recruited as part of a double-blind, randomized control study using a 1:1 ratio with parallel design (March 2022–February 2024). The MDD sample was recruited as part of an open-label study (February 2020–October 2024). Both studies took place within the University of Pennsylvania Department of Psychiatry with oversight by the University of Pennsylvania Institutional Review Board. All participants provided written informed consent.

Inclusion criteria were a primary diagnosis of BD or MDD; Montgomery-Åsberg Depression Rating Scale (MADRS)(Montgomery & Asberg, 1979) score ≥ 20; ≥ two prior failed treatments by Antidepressant Treatment History Form (ATHF) (Sackeim et al., 2019) criteria; on a stable medication regimen ≥ four weeks prior to initiating aiTBS treatment (including mood stabilizer for BD sample); and ages 22–70 years (BD sample) or 18–70 years (MDD sample). Exclusion criteria were a primary psychiatric diagnosis other than BD or MDD; rapid cycling with > four episodes per year (BD sample); MRI contraindication; pregnancy; diagnosis of stroke, epilepsy, or other neurological disorder; and current substance use disorder in the last three months.

### Participants

Fifty-four participants (34 BD, 20 MDD) were recruited from the University of Pennsylvania and the surrounding community (Figure S1). Of these 54, 17 failed screening (eight BD, nine MDD) and three withdrew (two BD, one MDD), leaving 34 participants (24 BD, 10 MDD) who completed the study. Of the remaining 24 participants with BD, 12 were randomized to the active aiTBS group and 12 to the sham aiTBS group. All ten participants with MDD received active aiTBS.

### aiTBS Procedure

Participants with BD were randomly assigned to receive either active or sham aiTBS by a statistician using permuted blocks allocation. Clinical assessors and treatment providers were separate individuals for all primary outcome measures. Participants, clinical assessors, and treatment providers were blinded. An analysis of the integrity of the blind was described previously (Sheline et al., 2024).

All BD and MDD participants completed ten sessions per day (one session per hour; 18,000 pulses per day) of active or sham imaging-guided aiTBS at 90% resting motor threshold for five days. Resting motor threshold was determined from electromyography recordings of the first dorsal interosseous muscle using the adaptive Parameter Estimation by Sequential Testing (PEST) algorithm (Mishory et al., 2004). A MagVenture (Farum, Denmark) MagPro X100 system with a double-sided Cool-B65 A/P coil was used, with sham and active sides labeled “A” or “B.” Sham treatment involved electrical pulses administered to mimic the sensation of receiving aiTBS. This approach enabled us to include a convincing sham condition without introducing electrically or magnetically induced changes to the electric field (e-field) at a biologically active level at the cortex (Balderston et al., 2022; Balderston, Beydler, et al., 2020; Balderston, Roberts, et al., 2020; Sheline et al., 2024), as described previously (e.g., Rastogi et al, 2017 (Rastogi et al., 2017)). Brainsight (Rogue Research, Montreal, Canada) neuronavigation software was used for TMS coil positioning (https://www.rogue-research.com/tms/brainsight-tms/).

### Imaging and fMRI Preprocessing

Participants completed MR scanning sessions at the University of Pennsylvania before and immediately (i.e., one day) after receiving aiTBS (Siemens 3T Magnetom Prisma Fit; 64-channel head coil; Erlangen, Germany). Sequences were rigorously tested as part of the Human Connectome Project (Van Essen et al., 2013) and optimized for Siemens Prisma scanners. During each scanning session, participants were instructed to keep their eyes on a fixation cross while BOLD resting-state fMRI data were acquired (gradient-echo EPI, 2 mm isotropic resolution, 52° flip angle, 800/37 ms TR/TE). T1- and T2*-weighted structural images were acquired for functional volume registration (T1w TurboFLASH, 0.9 x 0.9 x 1 mm resolution, 8° flip angle, 2200/4.67 ms TR/TE; T2*w SPACE, 0.9 x 0.9 x 1 mm resolution, variable flip angle, 3200/563 ms TR/TE). For the BD sample and the four final participants in the MDD sample, 23 minutes of BOLD resting-state fMRI data were acquired with pepolar field maps. For the first six participants in the MDD sample, eight minutes of BOLD resting-state fMRI data were acquired.

Images were preprocessed with fMRIPrep version 20.0.6 (Esteban et al., 2019) and xcpEngine version 1.1.0 (Ciric et al., 2018) using the fc-36p_despike preconfigured pipeline (https://github.com/PennLINC/xcpEngine/blob/master/designs/fc-36p_despike.dsn). Brain tissue was extracted and segmented from anatomical images and normalized to Montreal Neurological Institute (MNI; Grabner et al., 2006) standard space. BOLD single-band reference images were corrected for head motion using six-parameter rigid body realignment and registered to corresponding T1-weighted images with an affine transformation using the BBR cost function (Greve & Fischl, 2009). When present, pepolar field maps were used for susceptibility distortion correction. BOLD timeseries data were resampled into MNI standard space. Images were despiked with a scrubbing threshold of ∼0.13 mm; volumes with framewise displacement greater than 0.13 mm were removed. Confounds were regressed out. Frequencies were bandpass filtered with a 0.01–0.08 Hz passband, voxelwise regional homogeneity was calculated, and spatial smoothing was performed using a 6 mm FWHM SUSAN kernel (Smith & Brady, 1997).

### Target Generation

An individualized left dorsolateral prefrontal cortex (L dlPFC) stimulation target was generated for each participant based on their baseline MR data (Figure 1). In the BD sample, this was accomplished using the Cash et al. cluster method (Cash et al., 2021). Specifically, a network map using the subgenual anterior cingulate cortex (sgACC) as a seed was used as a proxy for the sgACC to improve the signal-to-noise ratio and reproducibility (Elbau et al., 2023; Li et al., 2014; Williams et al., 2018). The L dlPFC was excluded from this sgACC network map and timeseries values within the network map were correlated with those within the L dlPFC. The top 0.5% most anticorrelated voxels within the L dlPFC were then spatially clustered. The center of gravity of the largest resulting cluster was selected as the aiTBS target. Targeting was followed by e-field modeling (Balderston et al., 2022) to optimize individualized coil orientation and maximize e-field amplitude at the site of stimulation, as described in the Supplement.

In the MDD sample, targeting for the first seven participants utilized a mask of the L dlPFC retrieved from Neurosynth (Yarkoni et al., 2011), a functionally derived atlas that synthesizes functional connectivity results across thousands of studies to generate ROIs. This Neurosynth L dlPFC mask was then registered to the participant’s native space using FSL (Jenkinson et al., 2012) FLIRT with 12 degrees of freedom. A central location falling on a gyrus was selected from within this mask and used as the aiTBS target. For the final three MDD participants, the aiTBS target was defined as the region within the L dlPFC maximally anticorrelated with the sgACC. As in the BD sample, e-field modeling was then applied to optimize coil orientation for these functionally derived targets.

### Connectivity Calculations

The seven-network parcellation from Yeo et al. (Yeo et al., 2011) was used to define the DMN. Time series values were extracted from within the DMN. For each scanning session (before and one day after aiTBS), averaged voxelwise *Z*-normalized connectivity was calculated by applying the Global Brain Connectivity (GBC) protocol (Cole et al., 2010) to the voxels within the DMN. The GBC protocol calculates connectivity across all voxels within a given network simultaneously by computing average connectivity strength, producing an unbiased approach to the dysconnectivity location. As a secondary analysis, the GBC protocol was also utilized to determine whole-brain connectivity to ensure DMN results did not simply reflect a global change in connectivity.

### Clinical Assessments

The MADRS was used to assess the relationship between changes in depression severity and changes in connectivity. MADRS scores were collected before aiTBS, on each of the five treatment days, immediately following aiTBS (the day after the fifth treatment day), two weeks post-aiTBS (MDD sample only), four weeks post-aiTBS, and 12 weeks post-aiTBS (MDD sample only). Changes in other clinical measures were recorded for secondary analyses, as described in the Supplement. These measures included the Ruminative Thought Scale (RTS) (Brinker & Dozois, 2009) to assess rumination, the Scale for Suicidal Ideation (SSI) (Beck et al., 1979) to assess suicidality, the Insomnia Severity Index (ISI) (Bastien et al., 2001) to assess insomnia, and the Beck Depression Inventory–II (BDI) (Beck et al., 1996), a self-report depression scale, to corroborate depression improvement.

Current and previous medication trials were collected using ATHF criteria. Psychiatric diagnoses and comorbidities were corroborated using the Structured Clinical Interview for DSM-5 (SCID-5) (First et al., 2016); medical diagnoses were coded in accordance with the 10^th^ revision of the International Classification of Diseases (ICD-10) (Organization, 1993). Race, ethnicity, gender, sex assigned at birth, years of education, current medication, number of prior failed antidepressant trials, duration of primary diagnosis, and BD sample treatment allocation guesses were recorded; within the BD sample, these measures, along with within-group changes and between-group differences in MADRS scores, were reported previously (Sheline et al., 2024).

### Statistical Analysis

Analyses were performed using R version 4.3.2 (R Foundation for Statistical Computing, Vienna, Austria). Hypothesis tests were two-sided; *α* = .05. Baseline demographic characteristics were compared between groups (BD active, BD sham, and MDD) using Chi-square tests and ANOVAs with Tukey’s HSD for post-hoc analyses. Among those who received active aiTBS, the effects of targeting method (BD using Cash et al. cluster method vs. MDD using anatomical method vs. MDD using functional method) on connectivity change and MADRS change following aiTBS were assessed using ANOVAs. Similar ANOVAs were constructed to assess the effects of MRI scan parameters (those used for the first six MDD sample participants vs. those used for the BD sample and final four MDD sample participants) on connectivity change. ANOVAs assessing the effects of diagnostic sample membership (BD active vs. MDD) on change in connectivity and change in MADRS following active aiTBS were used to determine whether data from the two active samples (BD active and MDD) could be combined. Within each treatment group (combined active samples, BD active, MDD, and sham), paired *t*-tests were utilized to assess changes in *Z*-normalized connectivity, both within the DMN and across the whole brain, before versus after aiTBS. Linear mixed-effects models (LMEs) with degrees of freedom approximated using the Kenward-Roger method were used to assess the effects of study visit, active vs. sham aiTBS condition, their interaction, and MDD vs. BD diagnosis, with a random intercept for participants, on repeated scores from MADRS and other clinical measures. Pearson correlation coefficients were generated to assess the relationships between change in within-DMN connectivity and change in MADRS before versus after aiTBS among participants in each treatment group. Correlations between change in connectivity and change in MADRS scores were compared between active/sham aiTBS conditions using the Fisher *r*-to-*Z* transformation. A linear model was constructed to determine whether the relationship between change in connectivity and change in MADRS in the active group remained when controlling for differences in targeting methods, MRI scan parameters, and MDD vs. BD diagnosis. As a secondary analysis, an additional linear model was constructed to test whether treatment response, defined as a > 50% reduction in MADRS following active aiTBS, predicted change in within-DMN connectivity when controlling for diagnosis.

## RESULTS

### Participants

Thirty-four participants completed the study (*n* = 24 BD, *n* = 10 MDD). Among the 24 participants with BD, 12 were randomized to the active treatment group (6 female, 6 male; mean ± SD age, 43 ± 15.2 years) and 12 were randomized to the sham treatment group (6 female, 6 male; mean ± SD age, 44 ± 19.2 years). All ten participants in the MDD sample (6 female, 4 male; mean ± SD age, 39 ± 15.3 years) received active aiTBS. Detailed BD sample demographic information—including race, years of education, psychiatric and medical diagnoses, current medication, and prior antidepressant trials—was reported previously(Sheline et al., 2024). Demographic information for BD and MDD samples is reported in Table 1; there were no significant differences between groups (BD active, BD sham, and MDD) in any baseline variable except for number of prior depressive episodes (*F*_2,31_ = 4.26, *p* = .02), which Tukey’s post-hoc comparison revealed was lower in the active BD sample (mean ± SD, 3.3 ± 1.23) than in the MDD sample (mean ± SD, 4.9 ± 1.52; *p* = .03). Number of prior depressive episodes did not differ between those in the BD active sample and those in the BD sham sample (mean ± SD, 4.5 ± 1.24; *p* = .10) or between those in the MDD sample and those in the BD sham sample (*p* = .76). Adverse events, which did not differ in incidence between active and sham aiTBS recipients, are reported in the Supplement.

**Table 1.**
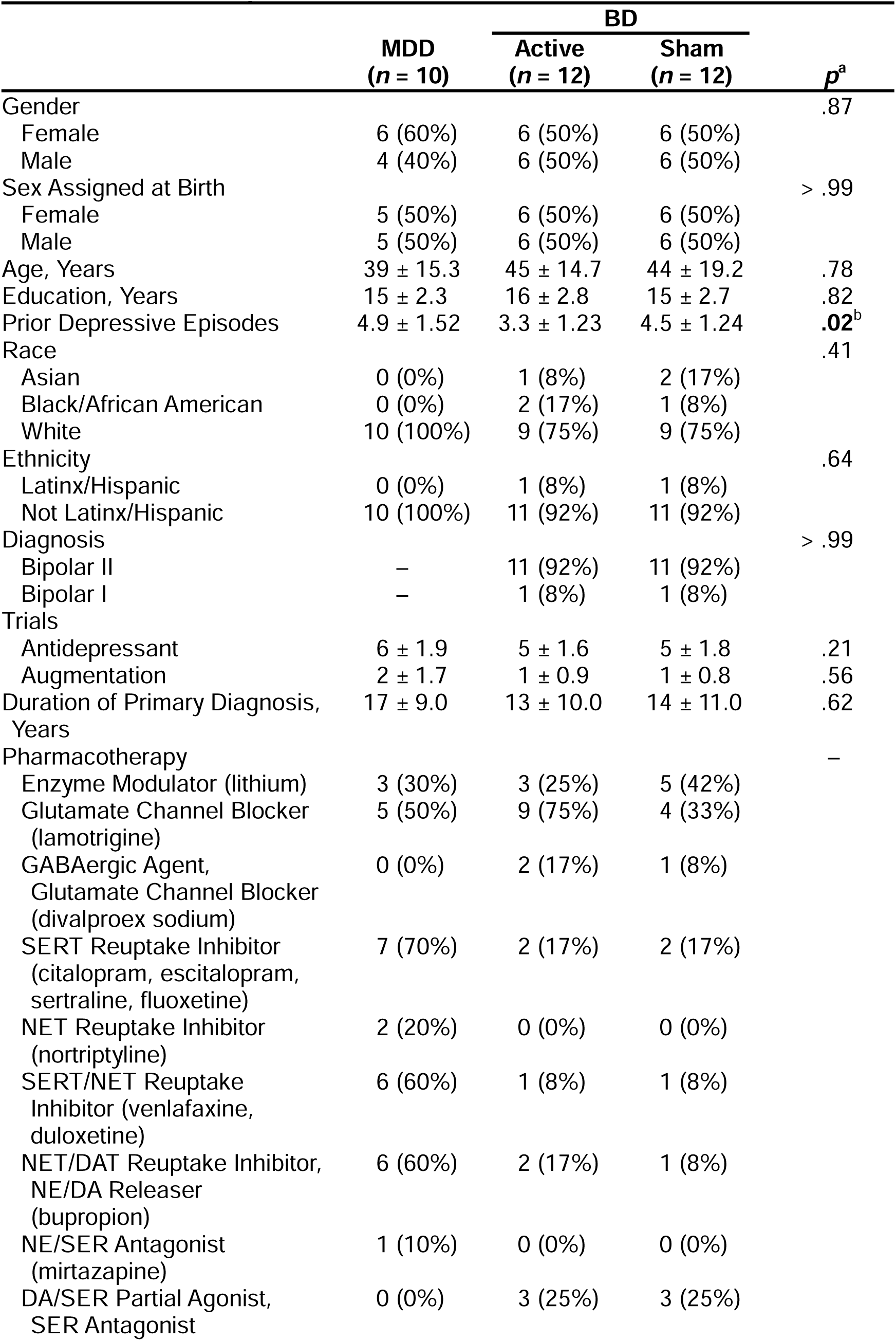
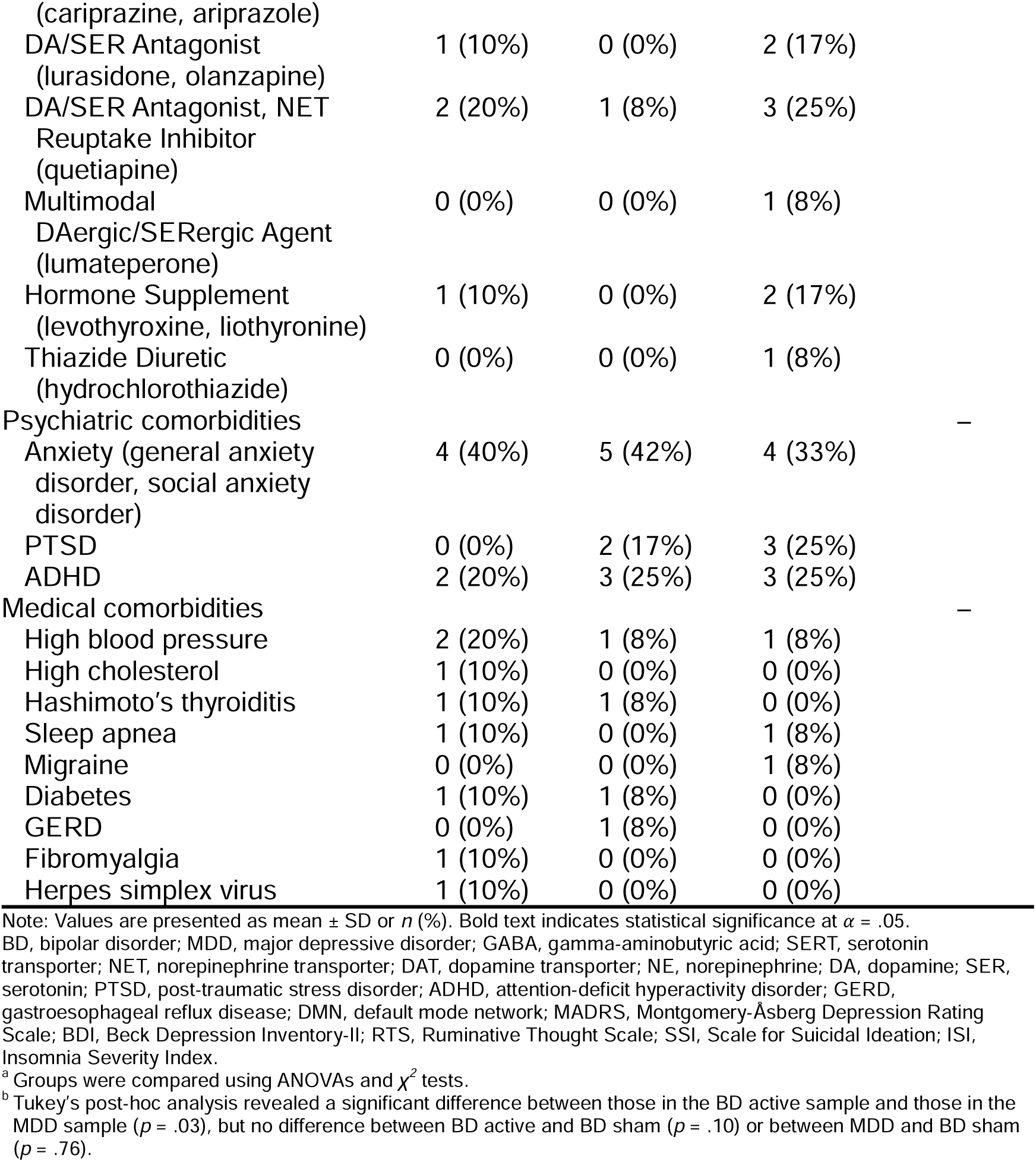
Baseline Sample Characteristics.

### Effects of Sample Differences

Among those who received active aiTBS, neither targeting method (Δ connectivity, *F*_1,20_ = 0.001, *p* = .98; Δ MADRS, *F*_1,20_ = 3.05, *p* = .10) nor BD vs. MDD diagnosis (Δ connectivity, *F*_1,20_ = 0.24, *p* = .63; Δ MADRS, *F*_1,20_ = 1.29, *p* = .27) had an effect on either of the primary outcome measures. Likewise, scanning parameters did not have an effect on change in connectivity (*F*_1,20_ = 0.10, *p* = .76). Therefore, active BD and MDD sample data were combined (*n* = 22 active aiTBS) for all following analyses; separate data for each sample are reported in Table S1.

### Connectivity Changes

Primary outcomes are reported in Table 2. Following treatment, combined BD/MDD active aiTBS group within-DMN connectivity was lower than at baseline (Δ mean ± SD *Z*-normalized Pearson’s *r* [*r_z_*], –0.017 ± 0.0270.; *t*[21] = –2.89, *p* = .01; Cohen’s *d* = –0.62; Figure 2). Changes in within-DMN connectivity were not different between active group BD and MDD diagnostic samples (*t*[14.62] = 0.47, *p* = .64; Cohen’s *d* = 0.21; Figure 3A). In the sham BD sample, there were no changes in within-DMN connectivity (Δ mean ± SD *r_z_*, –0.009 ± 0.0180; *t*[11] = –1.69, *p* = .12; Cohen’s *d* = –0.49) following sham aiTBS. Whole-brain connectivity did not change in either group, as reported in the Supplement.

**Table 2.**
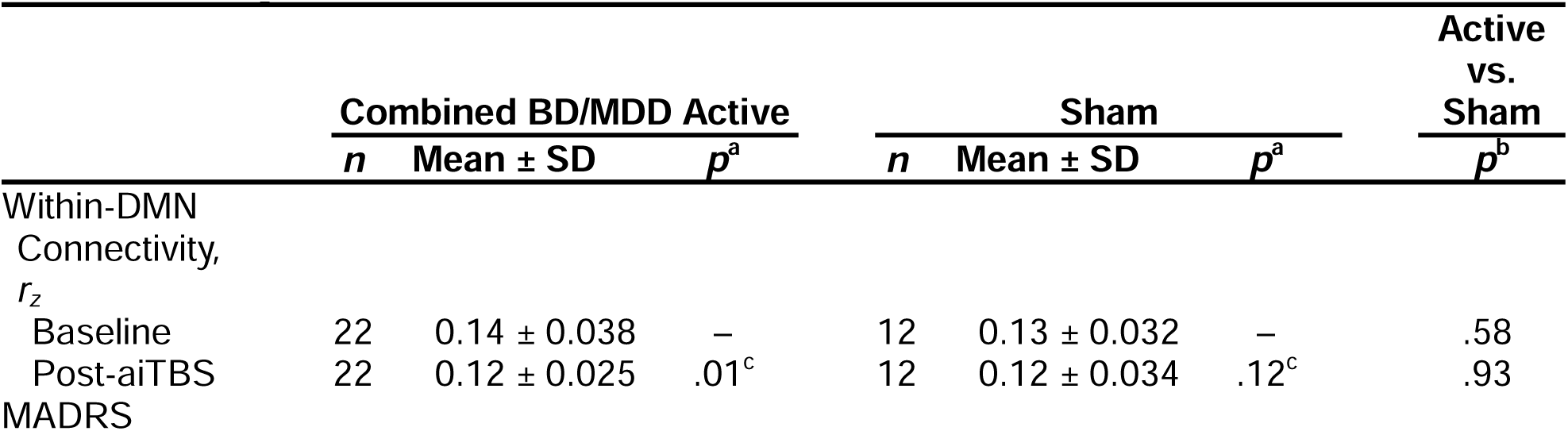
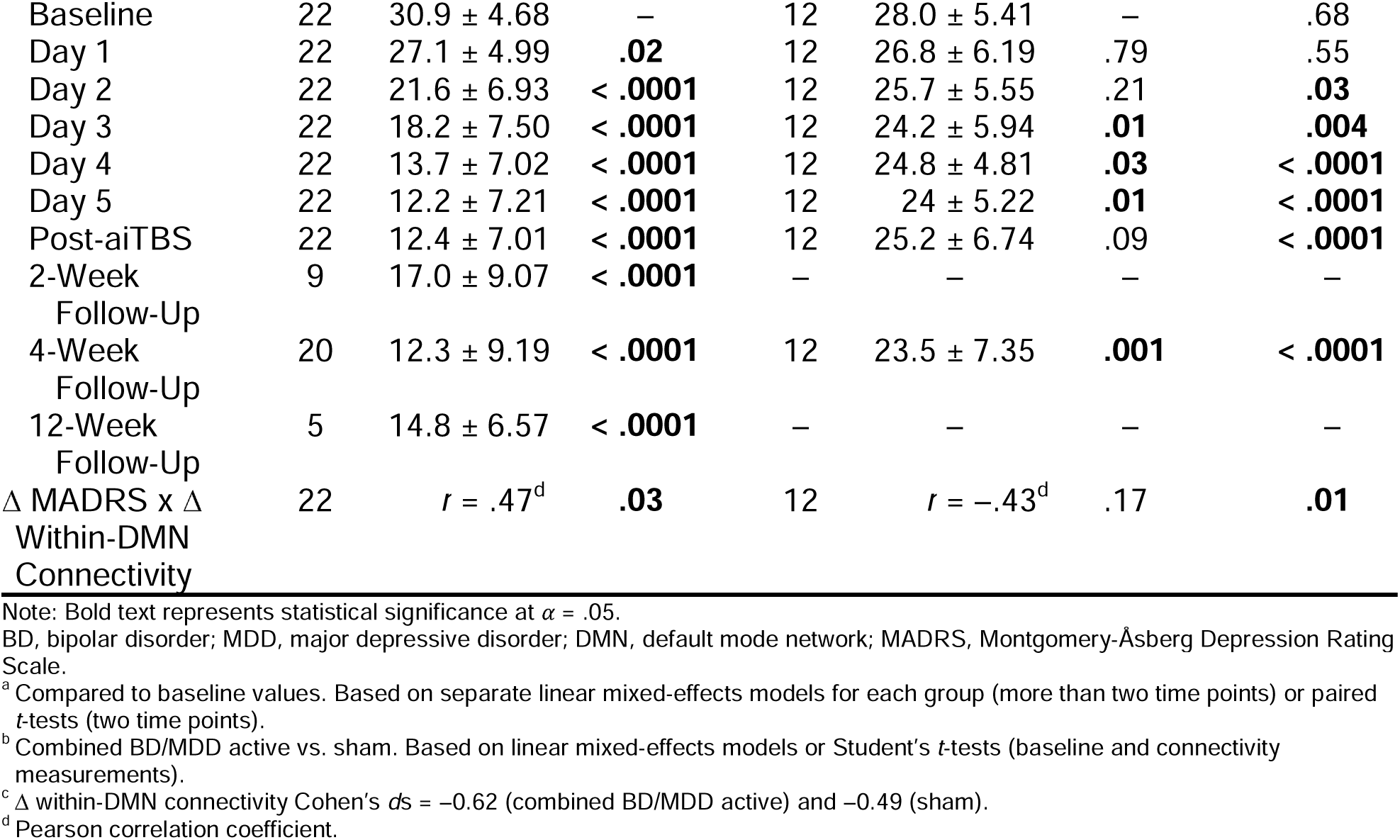
Primary Outcomes.

|  | Combined BD/MDD Active |  |  | Sham |  |  | Active vs. Sham |
| --- | --- | --- | --- | --- | --- | --- | --- |
| | $n$ | Mean ± SD | $p^a$ | $n$ | Mean ± SD | $p^a$ | $p^b$ |
| Within-DMN |  |  |  |  |  |  |  |
| Connectivity, |  |  |  |  |  |  |  |
| $r_z$ | | | | | | | |
| Baseline | 22 | 0.14 ± 0.038 | — | 12 | 0.13 ± 0.032 | — | .58 |
| Post-aiTBS | 22 | 0.12 ± 0.025 | .01<sup>c</sup> | 12 | 0.12 ± 0.034 | .12<sup>c</sup> | .93 |
| MADRS |  |  |  |  |  |  |  |
| Baseline | 22 | 30.9 ± 4.68 | – | 12 | 28.0 ± 5.41 | – | .68 |
| Day 1 | 22 | 27.1 ± 4.99 | **.02** | 12 | 26.8 ± 6.19 | .79 | .55 |
| Day 2 | 22 | 21.6 ± 6.93 | **< .0001** | 12 | 25.7 ± 5.55 | .21 | **.03** |
| Day 3 | 22 | 18.2 ± 7.50 | **< .0001** | 12 | 24.2 ± 5.94 | **.01** | **.004** |
| Day 4 | 22 | 13.7 ± 7.02 | **< .0001** | 12 | 24.8 ± 4.81 | **.03** | **< .0001** |
| Day 5 | 22 | 12.2 ± 7.21 | **< .0001** | 12 | 24 ± 5.22 | **.01** | **< .0001** |
| Post-aiTBS | 22 | 12.4 ± 7.01 | **< .0001** | 12 | 25.2 ± 6.74 | .09 | **< .0001** |
| 2-Week | 9 | 17.0 ± 9.07 | **< .0001** | – | – | – | – |
| Follow-Up |  |  |  |  |  |  |  |
| 4-Week | 20 | 12.3 ± 9.19 | **< .0001** | 12 | 23.5 ± 7.35 | **.001** | **< .0001** |
| Follow-Up |  |  |  |  |  |  |  |
| 12-Week | 5 | 14.8 ± 6.57 | **< .0001** | – | – | – | – |
| Follow-Up |  |  |  |  |  |  |  |
| Δ MADRS x Δ | 22 | $r = .47^d$ | **.03** | 12 | $r = -.43^d$ | .17 | **.01** |
| Within-DMN |  |  |  |  |  |  |  |
| Connectivity |  |  |  |  |  |  |  |

Note: Bold text represents statistical significance at $\alpha = .05$ .
BD, bipolar disorder; MDD, major depressive disorder; DMN, default mode network; MADRS, Montgomery-Åsberg Depression Rating Scale.
<sup>a</sup> Compared to baseline values. Based on separate linear mixed-effects models for each group (more than two time points) or paired *t*-tests (two time points).
<sup>b</sup> Combined BD/MDD active vs. sham. Based on linear mixed-effects models or Student's *t*-tests (baseline and connectivity measurements).
<sup>c</sup> Δ within-DMN connectivity Cohen's *ds* = –0.62 (combined BD/MDD active) and –0.49 (sham).
<sup>d</sup> Pearson correlation coefficient.

### MADRS Correlations

As reported in Table S1, no outcome measures were different between samples at baseline. An LME incorporating all participants (BD active, MDD, and sham) revealed a significant interaction between study visit and active vs. sham aiTBS condition on MADRS scores (*F*_7,234.01_ = 20.74, *p* < .0001), such that there was a significantly greater decrease in MADRS scores over time among those in the combined BD/MDD active aiTBS group than in those receiving sham aiTBS (estimated difference immediately post-aiTBS = –14.68; *p* < .0001). Combined active aiTBS group post-treatment MADRS scores were significantly lower than at baseline (estimated difference, –18.50; *p* < .0001), while sham condition MADRS scores were not (estimated difference, –2.75; *p* = .29). Active group MADRS scores were lower than those of the sham group starting on day two of aiTBS (estimated difference = –5.87; *p* = .03) and remained lower throughout the remainder of the study (*p*s < .0001–.004). MDD vs. BD diagnosis did not have an effect on MADRS scores (estimated difference = –4.04; *F*_1,31.16_ = 2.85, *p* = .10; Figure 3B). Likewise, there were no differences in responder status between BD active and MDD. samples, as reported in the Supplement.

Change in MADRS correlated with change in within-DMN connectivity in the combined BD/MDD active aiTBS group (*r* = .47, *p* = .03; Figure 4) but not in the sham group (*r* = –.43, *p* = .17). Moreover, change in MADRS and change in within-DMN connectivity were significantly more correlated in the combined active aiTBS group than in the sham group (*z* = 2.41, *p* = .01). The relationship between change in MADRS and change in within-DMN connectivity in the active aiTBS group remained significant (*partial R^2^* = .23, *p* = .04) even when controlling for targeting method (*partial R^2^* = .002, *p* = .86), MRI scan parameters (*partial R^2^* = .04, *p* = .42), and diagnosis (*partial R^2^* = .001, *p* = .90). Secondary clinical outcomes (SSI, ISI, RTS, and BDI) are reported in the Supplement.

## DISCUSSION

The current study examined aiTBS-induced brain connectivity changes and corresponding clinical improvement using identical stimulation protocols across patients with MDD and BD. Our recent randomized trial in treatment-refractory BD depression demonstrated that five days of active aiTBS led to significant reductions in MADRS scores, with response and remission rates comparable to those observed with aiTBS in MDD (Sheline et al., 2024). We now report that our data from that double-blind, randomized aiTBS trial in BD showed significant reductions in within-DMN connectivity following five days of 18,000 pulses/day to individualized L dlPFC targets using neuronavigation and e-field modeling. Similar within-DMN connectivity changes were found in our open-label MDD trial. We did not find whole-brain connectivity changes, suggesting the within-DMN connectivity changes we observed were not global.

The samples were well matched demographically and all received the same stimulation protocol—five days of treatment with ten aiTBS treatments per day using neuronavigation. Our use of a network-level sgACC map for the BD sample allowed for more robust and reproducible targeting and connectivity measurement to promote candidate marker precision. Although we did not include a healthy control group but focused instead on changes in treatment response, our results are consistent with existing literature reporting increased baseline connectivity within the DMN in MDD (Liston et al., 2014; Misir et al., 2023; Zhou et al., 2020). More recently, findings of increased within-DMN connectivity have been extended to BD (Cui et al., 2025; Fan et al., 2025; Zhang et al., 2024). In a sample of over 3000 people with MDD, BD, and healthy controls, Zhang et al. (Zhang et al., 2025)found increased DMN connectivity with multiple other regions and networks in both BD and MDD patients compared to healthy controls, although some additional changes—e.g., increased DMN connectivity with the inferior frontal gyrus—were evident in MDD alone. Longitudinal studies in BD found that increased within-DMN connectivity was associated with suicidal ideation (Wu et al., 2025) and illness exacerbations more generally (Cui et al., 2025), which normalized with treatment. It is worth noting, however, that some prior research failed to find increases in within-DMN connectivity in BD: Using spectral dynamic causal modeling, Fan et al. found decreased effective connectivity in BD compared to controls (Fan et al., 2025). In a review paper of 51 studies of BD, Sun et al. found that BD was associated with reduced intrinsic connectivity within all major brain networks, including the DMN (Sun et al., 2025). That said, this decreased connectivity may reflect methods that pointed to reduced global connectivity rather than a specific DMN effect.

We found that within-DMN connectivity decreased following aiTBS in the combined BD/MDD active group and in the BD active sample alone; however, change in within-DMN connectivity approached, but did not reach, significance in the MDD sample alone. This is likely due to the small sample size of this pilot study, which limits power, as evidenced both by the large combined active group effect (*d* = –0.62) and by the similarity between the BD active sample effect size (*d* = –0.59) and that of a recent Stanford Neuromodulation Therapy protocol reporting decreased within-DMN connectivity (*d* = –0.56) following stimulation (Batail et al., 2023), although their results did not survive Bonferroni correction. Given that diagnosis did not have an effect on within-DMN connectivity change, that within-DMN connectivity change was not different between the two active samples (MDD and BD active), and that the relationship between connectivity change and MADRS change remained significant when controlling for covariates including diagnosis, we believe the evidence suggests that effects were likely not driven by diagnostic group differences. Further, our finding that, despite our smaller sample size, the correlation between reduced within-DMN connectivity and reduced depressive symptoms following aiTBS is significantly greater in the active group than in the sham group also supports the significance of the effect and underscores the role of the DMN in depressive symptoms. Since a binary treatment response variable did not predict within-DMN connectivity change, the robust relationship between connectivity change and MADRS change suggests gradation in treatment response; alternatively, there could simply be inadequate power for a binary analysis—only four participants in each diagnostic sample did not respond to active aiTBS. Future research should attempt to replicate our findings with a larger sample.

Our results corroborate prior research identifying DMN changes associated with treatment response (Baeken et al., 2019; Chen et al., 2021; Fox et al., 2012; Liston et al., 2014; Weigand et al., 2018). Studies in MDD have found reductions in DMN activity with different treatment modalities, including antidepressant medication (Chin Fatt et al., 2020), electroconvulsive therapy (Moreno-Ortega et al., 2019), and TMS (Liston et al., 2014), including aiTBS (Gajawelli et al., 2024). A large meta-analysis of resting-state fMRI in MDD (*N* = 892) identified increased within-DMN connectivity at baseline and supported its predictive utility for treatment outcome (Zhou et al., 2020). Our finding that change in within-DMN connectivity correlated with change in depressive symptoms in the combined active aiTBS group, even when controlling for variation between samples, suggests that within-DMN connectivity may function as a candidate transdiagnostic marker and that a mechanism of aiTBS-induced reduction in depression symptoms may be targeting and reducing within-DMN connectivity. That said, this correlation was not evident in the active BD group alone. Because both within-DMN connectivity and MADRS scores decreased following aiTBS in the BD group, future research should attempt to replicate this result with a larger sample size as well as identify whether connectivity changes in individual regions within the DMN could be driving symptom improvement. Additionally, whether the greater number of prior depressive episodes in the MDD sample relates to the correlation between symptom improvement and reduced within-DMN connectivity should be explored.

Since rumination is a key activity of the DMN in depression (Geurts et al., 2020; Hasani et al., 2025; Lubbers et al., 2024; Sheline et al., 2009), the validity of within-DMN connectivity as a candidate transdiagnostic treatment marker is supported by our finding that aiTBS also reduced RTS scores—albeit only in the BD sample, despite extensive literature supporting this association in depression more generally (Misir et al., 2023; Zhou et al., 2020). This discrepancy between diagnostic samples may be due to differences in data collection time points; RTS scores weren’t significantly reduced in the BD sample until four weeks post-TMS, while RTS wasn’t administered at the four-week follow-up visit in the MDD sample, suggesting that it could take time for the rumination reduction to develop. This explanation is supported by the lack of a significant effect of MDD vs. BD diagnosis on RTS scores. Similarly, the significant effect of MDD vs. BD diagnosis on overall ISI scores could relate to data collection time points, given that ISI was not administered four weeks post-aiTBS in the MDD sample or on aiTBS day two in the BD sample. Overall MDD sample ISI scores could be higher because ISI data were on average collected earlier in the treatment course in the MDD sample, or alternatively, because insomnia may be more prevalent among those with MDD than those with BD (Guerrera et al., 2024). Future research should examine longitudinal RTS and ISI changes post-aiTBS across diagnostic samples to determine whether differences in RTS and ISI reduction are related to diagnosis or simply a function of time.

Finally, most (22/24) of our BD sample had a diagnosis of BD-II, which has been shown to sort genetically with MDD in genome-wide association studies (Zhou et al., 2020), structural equation modeling (Grotzinger et al., 2019), and twin studies (McGuffin et al., 2003). Whether our findings generalize to BD-I remains to be determined, as does whether reduction in within-DMN connectivity correlates with depressive symptom improvement in disorders with comorbid depression, such as post-traumatic stress disorder. Additionally, our study is limited by a relatively small sample size as well as heterogenous targeting methods and MRI acquisition parameters (8 vs. 23 minutes of resting-state data per scan and the use of pepolar field maps) resulting from utilizing samples collected in different experiments. Our findings should therefore be replicated in a larger study with consistent parameters. Future studies should explore the influence of neural circuit heterogeneity, medication effects, targeting methodology, and illness phase. Nonetheless, we are encouraged that despite these limitations, reduced within-DMN connectivity appears to be a candidate transdiagnostic network marker for depressive symptom reduction across MDD and BD.

## CRediT AUTHORSHIP CONTRIBUTION STATEMENT

**Yvette I. Sheline:** designed the study, obtained funding, evaluated participants for inclusion and exclusion criteria, supervised TMS treatment, rated participants for primary outcome measures, wrote the first draft, and edited subsequent drafts. **Walid Makhoul:** recruited and screened participants, obtained MRI scans pre- and post-treatment, conducted TMS treatment, conducted daily symptom measures, contributed to the manuscript, and edited the manuscript. **Alexandra S. Batzdorf:** conducted data analysis for primary and secondary outcome measures, identified targets for stimulation, contributed to the manuscript, created figures and tables, and edited the manuscript. **Kevin G. Lynch:** randomized the subjects, broke the blind, oversaw all the data analysis, and edited the manuscript. **Nicholas L. Balderston:** conducted e-field modeling, assisted with target determination, and edited the manuscript. **Robin F.H. Cash:** provided code for personalized targeting and edited the manuscript. **Conor M. Liston:** edited the manuscript. Dr. Sheline had full access to all the data in the study and takes responsibility for the integrity of the data and the accuracy of the data analysis.

## ACKNOWLEDGEMENTS

We wish to acknowledge our funding source, the Milken Institute. The Milken Institute had no role in the study design, data collection, data analysis, data interpretation, or writing of this report. We would like to thank Maria Prociuk for her assistance with manuscript preparation. We thank Dr. Claudia Baldassano, Director, Outpatient Bipolar Clinic, University of Pennsylvania for her assistance in referring patients with bipolar disorder. We thank Dr. Michael Thase for serving in the role of clinical monitor. We wish to thank all of our participants for their help in making this project a success.

Data from the BD sample were published previously in Sheline YI, Makhoul W, Batzdorf AS, Nitchie FJ, Lynch KG, Cash R, et al. (2024): Accelerated intermittent Theta-Burst Stimulation and treatment-refractory bipolar depression: A randomized clinical trial. JAMA Psychiatry. 81:936–941.

## DATA AVAILABILITY

Individual clinical data, including the individual neural targets (brain coordinates), will be made available upon publication of this manuscript. These data will be de-identified and uploaded to Mendeley Data.

---

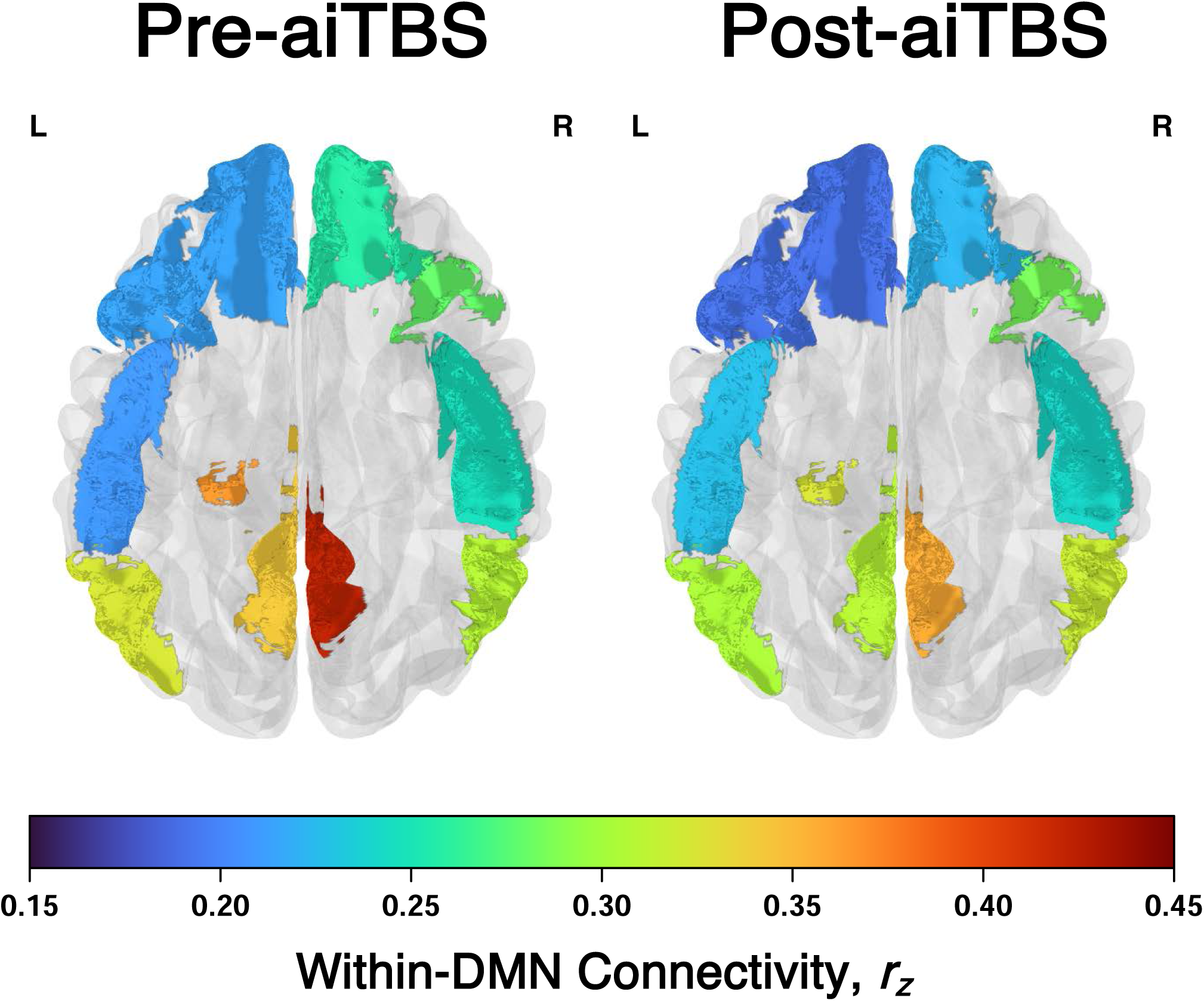

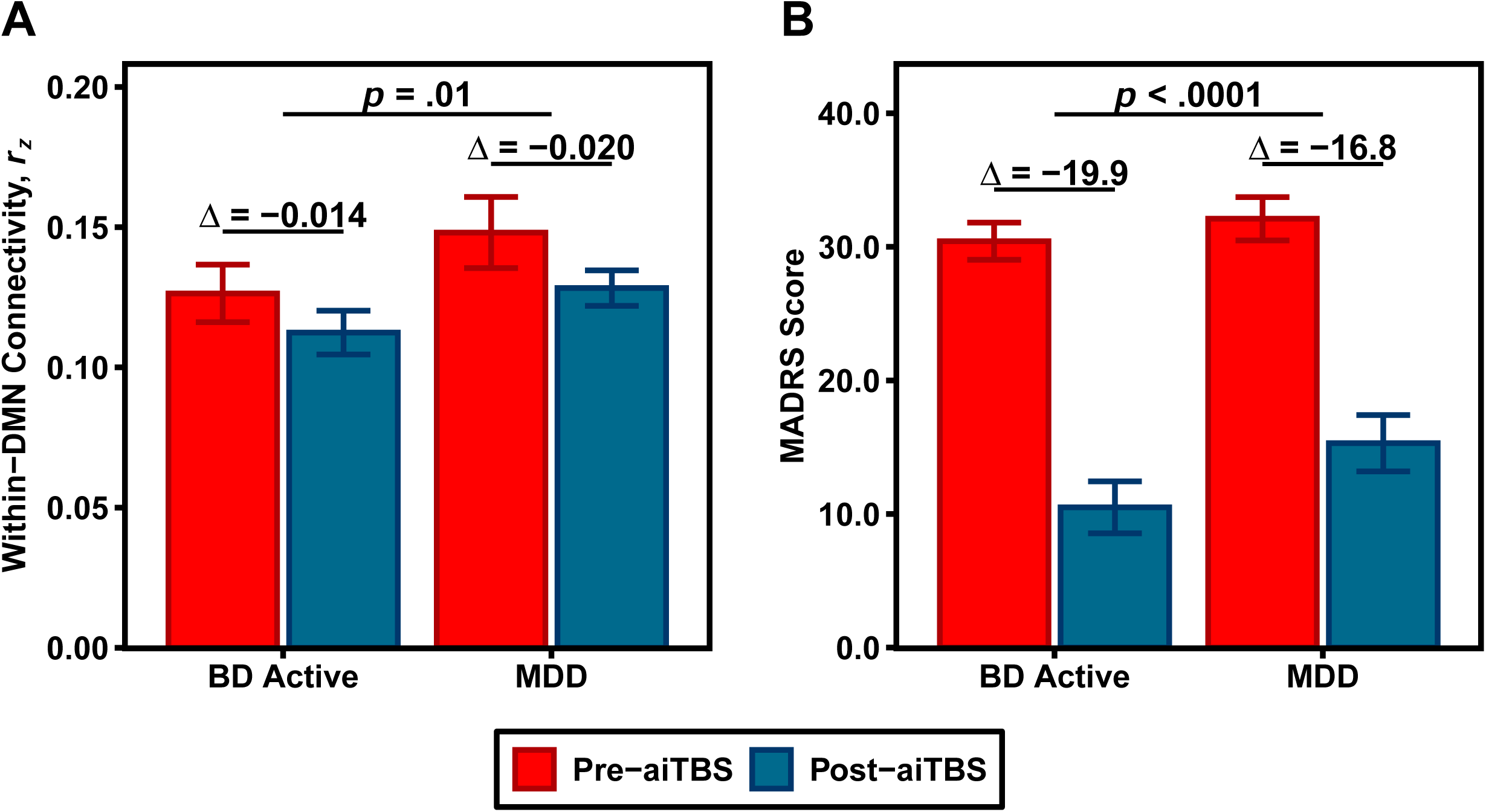

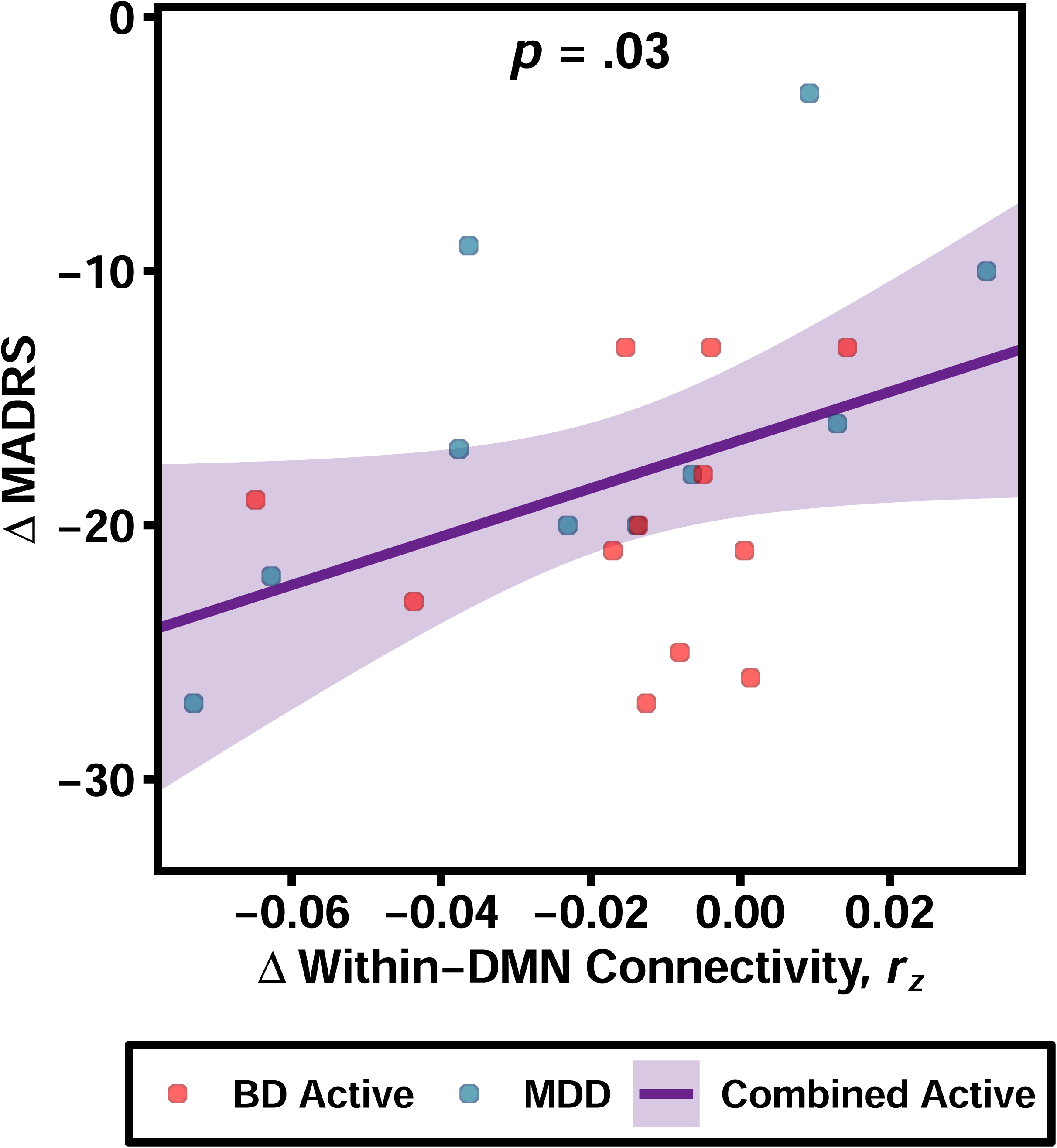

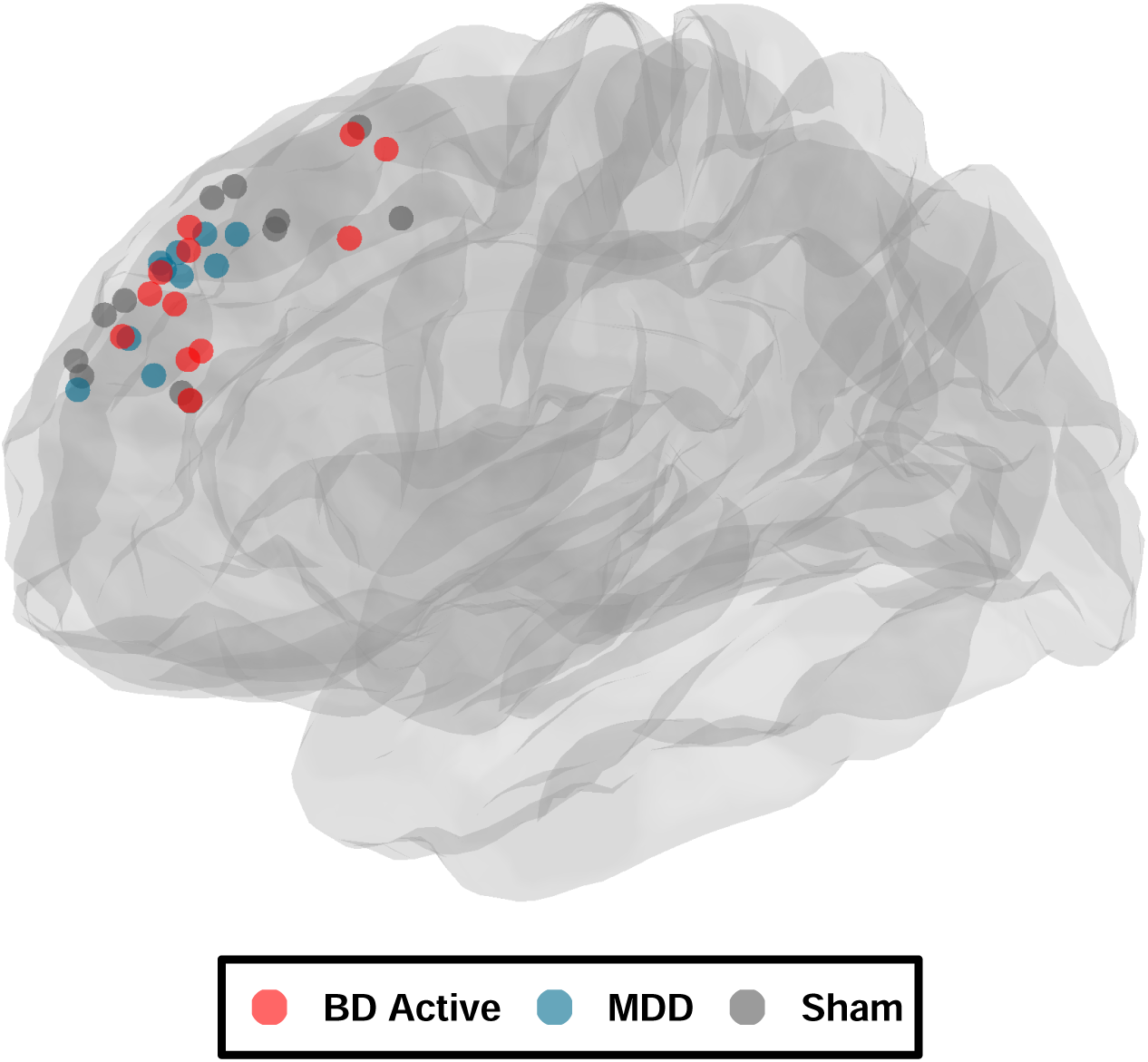

