## Supplementary material for "Decreased within-default mode network connectivity with accelerated intermittent theta burst stimulation across unipolar and bipolar depression: Candidate transdiagnostic circuit marker for treatment response": Methods and Materials

**Figure S1. CONSORT Flow Diagram**

Participant flow through the study. Adapted from Hopewell et al. (Hopewell et al., 2025).

**Major Depressive Disorder**

**Bipolar Disorder**

Enrollment

Assessed for eligibility

(*n* = 34)

Assessed for eligibility

(*n* = 20)

Excluded, did not meet inclusion criteria

(*n* = 8)

Excluded, did not meet inclusion criteria

(*n* = 9)

Randomized

(*n* = 26)

Allocation

Received active aiTBS

(*n* = 11)

Received active aiTBS

(*n* = 13)

Received sham aiTBS

(*n* = 13)

Withdrew due to time commitment

(*n* = 1)

Follow-Up

Withdrew due to time commitment

(*n* = 1)

Withdrew due to time commitment

(*n* = 1)

Analysis

Completed study; included in analyses

(*n* = 10)

Completed study; included in analyses

(*n* = 12)

Completed study; included in analyses

(*n* = 12)

**E-Field Modeling**

E-field modeling optimizes individualized coil orientation and maximizes e-field amplitude at the site of stimulation. This accounts for differences in the shape and orientation of dlPFC gyri coupled with electrical conductivity transitions between gray matter, white matter, and cerebrospinal fluid, which alter the current distribution dependent upon coil orientation (Thielscher et al., 2011). Head modeling and e-field calculations were performed using SimNIBS 2.1 (Puonti et al., 2020). Finite element models representing head and coil geometries were created for segmented tissue compartments (scalp, skull, cerebrospinal fluid, gray matter, and white matter) from the T1 (Thielscher et al., 2011) using gmsh subroutines (Thielscher et al., 2015). E-field models were conducted by projecting the target to the scalp using the knnsearch function in MATLAB. Twenty-four separate models were constructed at evenly spaced coil handle orientations tangential to the scalp. The resulting e-field maps (normE) were then sampled at the cortical target and the coil handle orientation corresponding to the maximal e-field was used during stimulation sessions (Balderston, Beydler, et al., 2020; Balderston, Roberts, et al., 2020).

**Secondary Clinical Measures**

The Ruminative Thought Scale (RTS; Brinker & Dozois, 2009) was used to assess rumination, the Scale for Suicidal Ideation (SSI; Beck et al., 1979) was used to assess suicidality, the Insomnia Severity Index (ISI; Bastien et al., 2001) was used to assess insomnia, and the Beck Depression Inventory–II (BDI; Beck et al., 1996), a self-report depression scale, was used to corroborate depression improvement. RTS was administered before aiTBS, on the second treatment day, the day immediately following aiTBS, and four weeks post-aiTBS (BD sample only); SSI was administered before aiTBS, on each of the five treatment days, one day immediately after aiTBS, two weeks post-aiTBS (MDD sample only), four weeks post-aiTBS, and 12 weeks post-aiTBS (MDD sample only); ISI was administered before aiTBS, on the first treatment day, on the second treatment day (MDD sample only), the day immediately following aiTBS, and four weeks post-aiTBS (BD sample only); and BDI was administered before aiTBS, on each of the five treatment days, the day immediately following aiTBS, two weeks post-aiTBS (MDD sample only), four weeks post-aiTBS, and 12 weeks post-aiTBS (MDD sample only).

**RESULTS**

**Adverse Events**

Adverse events included headache, dizziness, discomfort, and worsening of depressive symptoms. Headaches were reported by six (27.3%) active aiTBS recipients (BD active sample, *n* = 5 [41.7%]; MDD sample, *n* = 1 [10.0%]) and two (16.7%) sham group members. Dizziness was reported by one (4.6%) active aiTBS recipient (BD active sample, *n* = 1 [8.3%]; MDD sample, *n* = 0 [0%]) and no (0%) sham group members. Discomfort was reported by one (4.6%) active aiTBS recipient (BD active sample, *n* = 0 [0%]; MDD sample, *n* = 1 [8.3%]) and no (0%) sham group members. Likewise, worsening depressive symptoms were recorded in one (4.6%) active aiTBS recipient (BD active sample, *n* = 0 [0%]; MDD sample, *n* = 1 [8.3%]) and zero (0%) sham group members. There were no differences in incidence of any adverse event between combined BD/MDD active aiTBS and sham aiTBS groups (headache, *𝜒^2^* = 0.46, *df* = 1, *p* = .49; dizziness, *𝜒^2^* = 0.56, *df* = 1, *p* = .45; discomfort, *𝜒^2^* = 0.56, *df* = 1, *p* = .45; worsening depressive symptoms, *𝜒^2^* = 0.56, *df* = 1, *p* = .45) or between BD active, MDD, and sham samples (headache, *𝜒^2^* = 3.53, *df* = 2, *p* = .17; dizziness, *𝜒^2^* = 1.89, *df* = 2, *p* = .39; discomfort, *𝜒^2^* = 2.47, *df* = 2, *p* = .29; worsening depressive symptoms, *𝜒^2^* = 2.47, *df* = 2, *p* = .29).

**Secondary Outcomes**

Whole-brain analyses revealed no global changes in connectivity following aiTBS treatment in both active samples combined (*t*[21] = 1.69, *p* = .11, *d* = 0.36), just the BD active sample (*t*[11] = 1.34, *p* = .21, *d* = 0.39), or just the MDD sample (*t*[9] = 1.14, *p* = .29, *d* = 0.36). Likewise, no global changes were found following sham aiTBS (*t*[11] = 0.53, *p* = .61, *d* = 0.15).

In total, 14/22 (63.6%) active aiTBS recipients responded to treatment (8/12 [66.7%] BD active; 6/10 [60.0%] MDD). Proportion of responders did not differ between BD active and MDD diagnostic samples (*χ^2^* = 0.11, *df* = 1, *p* = .75). Responder status did not have an effect on change in within-DMN connectivity (*F*_1,20_ = 0.26, *p* = .62).

There was a significant interaction between study visit and active vs. sham aiTBS condition on SSI scores (*F*_7,224.15_ = 2.06, *p* = .049), such that SSI scores decreased over time significantly more in the combined BD/MDD active aiTBS group than in the sham group (estimated difference immediately post-aiTBS = –4.49; *p* = .03). While active group SSI scores were lower than at baseline from aiTBS day three (estimated difference, –4.22; *p* = .0003) through the remainder of the study (*p*s < .0001–.04), sham group SSI scores were not (day three estimated difference, –1.92; *p*s = .13–.46). MDD vs. BD diagnosis did not have an effect on SSI scores (estimated difference = –2.15; *F*_1,31.21_ = 1.27, *p* = .27).

There was a significant study visit x active vs. sham aiTBS condition interaction on ISI scores (*F*_3,77.05_ = 3.93, *p* = .01); ISI scores became significantly lower over time in the combined BD/MDD active aiTBS group than in the sham group (estimated difference immediately post-aiTBS = –6.03; *p* = .02). Combined BD/MDD active group ISI scores were lower than at baseline on aiTBS day one (estimated difference = –2.45; *p* = .04), immediately post-aiTBS (estimated difference = –5.34; *p* < .0001), and four weeks later (estimated difference = –4.95; *p* < .0001), while sham aiTBS group ISI scores were not (estimated difference immediately post-aiTBS = –1.42; *p*s = .37–.67). Additionally, active group ISI scores were lower than those of the sham group on aiTBS day one (estimated difference = –5.42; *p* = .03), immediately post-aiTBS (estimated difference = –6.03; *p* = .02), and four weeks later (estimated difference = –5.75; *p* = .02). Although ISI scores were not different between samples at baseline and ISI scores decreased over time in both active BD and MDD samples, MDD vs. BD diagnosis had a significant effect on ISI scores, in that overall ISI scores were lower among those with BD than those with MDD (estimated difference = –6.25; *F*_1,29.19_ = 5.10, *p* = .03).

There was a significant interaction between study visit and active vs. sham aiTBS on BDI scores (*F*_7,229.06_ = 3.47, *p* = .002), such that BDI scores became lower over time significantly more in the combined BD/MDD active aiTBS group than in the sham group (estimated difference four weeks post-aiTBS = –10.15; *p* = .01). Combined BD/MDD active group BDI scores were lower than at baseline from aiTBS day one (estimated difference = –4.95; *p* = .03) through the remainder of the study (all *p*s < .0001). Similarly, sham group BDI scores were lower than at baseline from aiTBS day two (estimated difference = –7.75; *p* = .01) through the remainder of the study (*p*s = .0001–.001); however, active group BDI scores were lower than sham group BDI scores on aiTBS day four (estimated difference = –7.85; *p* = .04), on aiTBS day five (estimated difference = –8.93; *p* = .02), and four weeks post-aiTBS (estimated difference = –10.15; *p* = .01). MDD vs. BD diagnosis did not have an effect on BDI scores (estimated difference = –3.72; *F*_1,31.22_ = 1.27, *p* = .27).

When incorporating all participants, there was no interaction between study visit and active vs. sham aiTBS condition on RTS scores (*F*_3,76.11_ = 0.70, *p* = .55). Active aiTBS group RTS scores were lower than at baseline four weeks after aiTBS (estimated difference = –17.46; *p* = .01), while sham group RTS scores were not (estimated difference = –6.34; *p* = .58), but at four weeks post-aiTBS, active group RTS scores were not significantly lower than those of the sham group (estimated difference = –12.00; *p* = .30). When looking the MDD and BD active samples separately, there was no effect of study visit on RTS scores in the MDD group (*F*_2,9.11_ = 0.16, *p* = .86), but there was an effect in the BD active group (*F*_3,33.00_ = 4.28, *p* = .01). MDD sample RTS scores were not lower than at baseline on either the second treatment day (estimated difference = 3.89; *p* = .84) or immediately post-aiTBS (estimated difference = –0.33; *p* > .99). The RTS was not administered four weeks post-aiTBS in the MDD sample. As in the MDD sample, active BD sample RTS scores were not lower than at baseline on the second treatment day (estimated difference = –4.50; *p* = .74) or immediately post-aiTBS (estimated difference = –7.92; *p* = .37), but they were lower than at baseline four weeks post-aiTBS (estimated difference = –19.17; *p* = .01). MDD vs. BD diagnosis did not have a significant effect on RTS scores (estimated difference = –0.06; *F*_1,28.39_ < 0.01, *p* > .99).

| **Table S1. Outcomes for Each Sample** | | | | | | | | | | | | | | | | | | | | | | | | | | | |
| --- | --- | --- | --- | --- | --- | --- | --- | --- | --- | --- | --- | --- | --- | --- | --- | --- | --- | --- | --- | --- | --- | --- | --- | --- | --- | --- | --- |
|  |  | |  | | | | | | | |  |  | | | | | | | | | | | | |  | **Active vs. Sham** | |
|  |  | |  | | | | | | | |  | **BD** | | | | | | | | | | | | |  |  |  |
|  |  | | **MDD** | | | | | | | |  | **Active** | | | | | |  | **Sham** | | | | | |  |  |  |
|  |  | | ***n*** |  | **Mean** | | **± SD** | | ***p*^a^** | |  | ***n*** |  | **Mean** | **± SD** | ***p*^a^** | |  | ***n*** |  | **Mean** | **± SD** | ***p*^a^** | |  | ***p*^b^** | |
| **Primary Outcomes** | | | | | | | | | | | | | | | | | | | | | | | | | | | |
| Within-DMN Connectivity, *r_z_* |  | |  |  |  | |  | |  |  |  |  |  |  |  |  |  |  |  |  |  |  |  |  |  |  |  |
| Baseline |  | | 10 |  | 0.15 | | ± 0.040 | |  | – |  | 12 |  | 0.13 | ± 0.036 |  | – |  | 12 |  | 0.13 | ± 0.032 |  | – |  |  | .58 |
| Post-aiTBS |  | | 10 |  | 0.13 | | ± 0.020 | |  | .10^c^ |  | 12 |  | 0.11 | ± 0.027 |  | **.04**^c^ |  | 12 |  | 0.12 | ± 0.034 |  | .12^c^ |  |  | .93 |
| MADRS |  | |  |  |  | |  | |  |  |  |  |  |  |  |  |  |  |  |  |  |  |  |  |  |  |  |
| Baseline |  | | 10 |  | 31.5 | | ± 4.70 | |  | – |  | 12 |  | 30.4 | ± 4.81 |  | – |  | 12 |  | 28.0 | ± 5.41 |  | – |  |  | .68 |
| Day 1 |  | | 10 |  | 27.7 | | ± 4.85 | |  | .23 |  | 12 |  | 26.6 | ± 5.26 |  | .07 |  | 12 |  | 26.8 | ± 6.19 |  | .79 |  |  | .55 |
| Day 2 |  | | 10 |  | 23.3 | | ± 7.63 | |  | **.0002** |  | 12 |  | 20.2 | ± 6.27 | **<** | **.0001** |  | 12 |  | 25.7 | ± 5.55 |  | .21 |  |  | **.03** |
| Day 3 |  | | 10 |  | 20.5 | | ± 6.98 | | **<** | **.0001** |  | 12 |  | 16.2 | ± 7.65 | **<** | **.0001** |  | 12 |  | 24.2 | ± 5.94 |  | **.01** |  |  | **.004** |
| Day 4 |  | | 10 |  | 16.7 | | ± 5.76 | | **<** | **.0001** |  | 12 |  | 11.2 | ± 7.23 | **<** | **.0001** |  | 12 |  | 24.8 | ± 4.81 |  | **.03** |  | **<** | **.0001** |
| Day 5 |  | | 10 |  | 14.6 | | ± 6.29 | | **<** | **.0001** |  | 12 |  | 10.2 | ± 7.55 | **<** | **.0001** |  | 12 |  | 24 | ± 5.22 |  | **.01** |  | **<** | **.0001** |
| Post-aiTBS |  | | 10 |  | 14.7 | | ± 6.96 | | **<** | **.0001** |  | 12 |  | 10.5 | ± 6.74 | **<** | **.0001** |  | 12 |  | 25.2 | ± 6.74 |  | .09 |  | **<** | **.0001** |
| 2-Week Follow-Up |  | | 9 |  | 17.0 | | ± 9.07 | | **<** | **.0001** |  | – |  |  | – |  | – |  | – |  |  | – |  | – |  |  | – |
| 4-Week Follow-Up |  | | 8 |  | 17.9 | | ± 10.09 | | **<** | **.0001** |  | 12 |  | 8.6 | ± 6.60 | **<** | **.0001** |  | 12 |  | 23.5 | ± 7.35 |  | **.001** |  | **<** | **.0001** |
| 12-Week Follow-Up |  | | 5 |  | 14.8 | | ± 6.57 | | **<** | **.0001** |  | – |  |  | – |  | – |  | – |  |  | – |  | – |  |  | – |
| ∆ MADRS x ∆ Within-DMN Connectivity |  | | 10 |  | *r* | | = .70^d^ | |  | **.02** |  | 12 |  | *r* | = .15^d^ |  | .65 |  | 12 |  | *r* | = –.43^d^ |  | .17 |  |  | **.01** |
| **Secondary Outcomes** | | | | | | | | | | | | | | | | | | | | | | | | | | | |
| SSI |  | |  |  |  | |  | |  |  |  |  |  |  |  |  |  |  |  |  |  |  |  |  |  |  |  |
| Baseline |  | | 10 |  | 8.0 | | ± 6.16 | |  | – |  | 12 |  | 6.7 | ± 6.18 |  | – |  | 12 |  | 8.3 | ± 6.30 |  | – |  |  | .34 |
| Day 1 |  | | 10 |  | 6.3 | | ± 4.88 | |  | .47 |  | 10 |  | 8.3 | ± 6.45 |  | .88 |  | 12 |  | 7.6 | ± 6.53 |  | .94 |  |  | .42 |
| Day 2 |  | | 10 |  | 7.0 | | ± 5.46 | |  | .87 |  | 10 |  | 4.4 | ± 4.25 |  | .16 |  | 12 |  | 5.1 | ± 4.80 |  | **.01** |  |  | .70 |
| Day 3 |  | | 10 |  | 5.7 | | ± 4.85 | |  | .18 |  | 10 |  | 2.8 | ± 3.52 |  | **.01** |  | 12 |  | 6.3 | ± 5.69 |  | .27 |  |  | .10 |
| Day 4 |  | | 10 |  | 4.8 | | ± 4.69 | |  | **.02** |  | 10 |  | 2.6 | ± 2.88 |  | **.004** |  | 12 |  | 5.9 | ± 5.84 |  | .12 |  |  | .09 |
| Day 5 |  | | 10 |  | 4.4 | | ± 4.67 | |  | **.01** |  | 10 |  | 2.0 | ± 2.94 |  | **.001** |  | 12 |  | 6.2 | ± 5.41 |  | .20 |  |  | **.04** |
| Post-aiTBS |  | | 10 |  | 4.6 | | ± 4.93 | |  | **.01** |  | 12 |  | 1.8 | ± 3.16 |  | **.001** |  | 12 |  | 6.6 | ± 5.96 |  | .40 |  |  | **.04** |
| 2-Week Follow-Up |  | | 9 |  | 5.9 | | ± 5.01 | |  | .14 |  | – |  |  | – |  | – |  | – |  |  | – |  | – |  |  | – |
| 4-Week Follow-Up |  | | 8 |  | 3.8 | | ± 5.39 | |  | **.01** |  | 12 |  | 2.0 | ± 4.26 |  | **.002** |  | 12 |  | 5.8 | ± 5.52 |  | .10 |  |  | .07 |
| 12-Week Follow-Up |  | | 5 |  | 2.2 | | ± 2.39 | |  | **.01** |  | – |  |  | – |  | – |  | – |  |  | – |  | – |  |  | – |
| ISI |  | |  |  |  | |  | |  |  |  |  |  |  |  |  |  |  |  |  |  |  |  |  |  |  |  |
| Baseline |  | | 6 |  | 17.3 | | ± 5.39 | |  | – |  | 12 |  | 11.8 | ± 6.36 |  | – |  | 12 |  | 13.6 | ± 6.54 |  | – |  |  | .39 |
| Day 1 |  | | 1 |  | 12.0 | | – | |  | .52 |  | 12 |  | 8.8 | ± 5.71 |  | **.02** |  | 12 |  | 14.5 | ± 6.42 |  | .67 |  |  | **.03** |
| Day 2 |  | | 5 |  | 15.6 | | ± 4.56 | |  | .52 |  | – |  |  | – |  | – |  | – |  |  | – |  | – |  |  | – |
| Post-TMS |  | | 6 |  | 12.2 | | ± 4.02 | |  | **.01** |  | 12 |  | 6.3 | ± 5.26 | **<** | **.0001** |  | 12 |  | 12.2 | ± 6.58 |  | .37 |  |  | **.02** |
| 4-Week Follow-Up |  | | – |  |  | | – | |  | – |  | 12 |  | 6.6 | ± 5.11 | **<** | **.0001** |  | 12 |  | 12.3 | ± 6.07 |  | .46 |  |  | **.02** |
| BDI |  | |  |  |  | |  | |  |  |  |  |  |  |  |  |  |  |  |  |  |  |  |  |  |  |  |
| Baseline |  | | 10 |  | 29.5 | | ± 5.64 | |  | – |  | 12 |  | 30.4 | ± 9.90 |  | – |  | 12 |  | 29.0 | ± 12.26 |  | – |  |  | .85 |
| Day 1 |  | | 10 |  | 25.3 | | ± 6.41 | |  | .53 |  | 12 |  | 24.8 | ± 11.07 |  | **.04** |  | 12 |  | 23.3 | ± 12.15 |  | .10 |  | > | .99 |
| Day 2 |  | | 10 |  | 22.1 | | ± 8.48 | |  | .06 |  | 12 |  | 18.2 | ± 9.52 | **<** | **.0001** |  | 12 |  | 21.3 | ± 8.51 |  | .01 |  |  | .43 |
| Day 3 |  | | 10 |  | 18.5 | | ± 7.01 | |  | **.001** |  | 12 |  | 16.1 | ± 12.51 | **<** | **.0001** |  | 12 |  | 18.5 | ± 9.11 |  | .0002 |  |  | .42 |
| Day 4 |  | | 10 |  | 17.0 | | ± 9.31 | |  | **.0002** |  | 12 |  | 10.8 | ± 11.24 | **<** | **.0001** |  | 12 |  | 19.8 | ± 7.02 |  | .001 |  |  | **.04** |
| Day 5 |  | | 10 |  | 13.5 | | ± 6.77 | | **<** | **.0001** |  | 12 |  | 10.9 | ± 11.71 | **<** | **.0001** |  | 12 |  | 19.3 | ± 7.49 |  | .001 |  |  | **.02** |
| Post-aiTBS |  | | 10 |  | 19.9 | | ± 7.87 | |  | **.01** |  | 12 |  | 11.0 | ± 10.86 | **<** | **.0001** |  | 12 |  | 20.0 | ± 7.15 |  | .002 |  |  | .08 |
| 2-Week Follow-Up |  | | 8 |  | 12.9 | | ± 8.72 | | **<** | **.0001** |  | – |  |  | – |  | – |  | – |  |  | – |  | – |  |  | – |
| 4-Week Follow-Up |  | | 6 |  | 14.2 | | ± 11.82 | |  | **.0002** |  | 12 |  | 8.1 | ± 6.82 | **<** | **.0001** |  | 11 |  | 18.6 | ± 6.44 |  | **.001** |  |  | **.01** |
| 12-Week Follow-Up |  | | 4 |  | 12.5 | | ± 7.23 | |  | **.0003** |  | – |  |  | – |  | – |  | – |  |  | – |  | – |  |  | – |
| RTS |  | |  |  |  | |  | |  |  |  |  |  |  |  |  |  |  |  |  |  |  |  |  |  |  |  |
| Baseline |  | | 6 |  | 83.3 | | ± 16.85 | |  | – |  | 12 |  | 88.5 | ± 27.23 |  | – |  | 12 |  | 87.7 | ± 34.80 |  | – |  |  | .94 |
| Day 2 |  | | 5 |  | 88.8 | | ± 26.95 | |  | .84 |  | 12 |  | 84.0 | ± 26.08 |  | .74 |  | 12 |  | 87.2 | ± 32.01 | > | .99 |  |  | .84 |
| Post-TMS |  | | 6 |  | 83.0 | | ± 21.87 | | > | .99 |  | 12 |  | 80.6 | ± 27.60 |  | .37 |  | 12 |  | 85.1 | ± 28.92 |  | .93 |  |  | .75 |
| 4-Week Follow-Up |  | | – |  |  | | – | |  | – |  | 12 |  | 69.3 | ± 30.40 |  | **.01** |  | 11 |  | 81.6 | ± 26.08 |  | .64 |  |  | .31 |
| Whole-Brain Connectivity, *r_z_* | |  |  |  |  | |  | |  |  |  |  |  |  |  |  |  |  |  |  |  |  |  |  |  |  |  |
| Baseline |  | | 10 |  | | 0.004 | ± 0.003 |  | | – |  | 12 |  | 0.01 | ± 0.002 |  | – |  | 12 |  | 0.01 | ± 0.002 |  | – |  |  | .39 |
| Post-TMS |  | | 10 |  | 0.01 | | ± 0.003 | |  | .29^e^ |  | 12 |  | 0.01 | ± 0.002 |  | .21^e^ |  | 12 |  | 0.01 | ± 0.003 |  | .61^e^ |  |  | .63 |
| Note: Bold text represents statistical significance at 𝛼 = .05.  BD, bipolar disorder; MDD, major depressive disorder; DMN, default mode network; MADRS, Montgomery-Åsberg Depression Rating Scale; SSI, Scale for Suicidal Ideation; ISI, Insomnia Severity Index; BDI, Beck Depression Inventory–II; RTS, Ruminative Thought Scale.   a Compared to baseline values. Based on separate linear mixed-effects models for each sample (more than two time points) or paired *t*-tests (two time points). b When data exist from all samples, combined BD/MDD active vs. sham; otherwise, BD active vs. sham. Based on linear mixed-effects models or Student’s *t*-tests (baseline and connectivity measurements). c ∆ within-DMN connectivity Cohen’s *d*s = –0.59 (MDD), –0.66 (BD active), and –0.49 (sham). d Pearson correlation coefficient. e ∆ whole-brain connectivity Cohen’s *d*s = 0.36 (MDD), 0.39 (BD active), and 0.15 (sham). | | | | | | | | | | | | | | | | | | | | | | | | | | | |
